# Linking the pathway from social media health information seeking to health misinformation sharing: A moderated serial mediation model

**DOI:** 10.1101/2023.08.18.23294258

**Authors:** Chen Luo, Yulong Tang, Yuying Deng, Yuru Li

## Abstract

Seeking health information from social media has become prominent in recent years. Meanwhile, the proliferation of online health misinformation keeps abreast of this tendency and sparks grave concerns. Drawing upon the S-O-R (Stimulus-Organism-Response) model and the cognitive load theory, the current study aims to clarify the relationship between social media health information seeking and health misinformation sharing with a focus on the Chinese middle-aged or above group, which has been deemed susceptible to online misinformation. Results of structural equation modeling based on an online survey (*N* = 388) disclosed a serial mediation process with health information overload and misperceptions as sequential mediators. Interestingly, while health misperceptions were positively related to misinformation sharing intention, health information overload was not. Furthermore, as a critical information processing predisposition, the need for cognition only buffered the positive association between information seeking and information overload. Overall, besides proposing a moderated serial mediation model to better comprehend the psychological mechanism underlying health misinformation sharing, this study highlights the importance of zooming into the organism part and the necessity of distinguishing between information overload and misperceptions in the context of health misinformation. Theoretical implications for unraveling online health misinformation sharing and practical implications for boosting immunity against health misinformation among at-risk groups are discussed.

## 1. Introduction

Seeking health information from cyberspace has become increasingly prevalent (Zheng & Tandoc, 2022). Among all the online channels, social media platforms have garnered particular popularity due to their incomparable efficiency in health information creation, retrieval, and sharing (Zhao & Zhang, 2017). Albeit the virtues of social media in health information circulation, whether it serves as a blessing or a bane highly depends on information quality and the characteristics of information seekers (Wu et al., 2022; Zhang et al., 2021). In recent years, the menace of online health misinformation has aroused solemn concerns and heated discussions about possible coping strategies (Bode & Vraga, 2018; Kim et al., 2023; Oktavianus & Bautista, 2023). Defined as health information that lacks support from the best available medical evidence or medical expert consensus (Nyhan & Reifler, 2010; Tan et al., 2015), health misinformation proliferates on social media and induces a series of negative outcomes, such as adopting unscientific precautionary measures and fostering mistrust in public health institutions (Chou et al., 2018; Melki et al., 2023). Moreover, health misinformation is especially detrimental to specific age groups, such as the middle-aged or above, which is susceptible to online health misinformation but enthusiastic about sharing health information on social media (Guan, 2019; Wu et al., 2022).

The rampant social media health misinformation and its deleterious consequences call for an in-depth inquiry into why health misinformation has been shared in the social media context, especially given that relevant explanations are insufficiently developed (Apuke et al., 2022; Apuke & Omar, 2021; Chou, 2018; Wu & Pei, 2022). Sharing misinformation is worrisome because it undoubtedly amplifies the impact scope of misinformation and aggravates its undesirable ramifications (Laato et al., 2020; Tang et al., 2023). Therefore, the current study heeds the call by formulating a psychological mechanism to explicate why the middle-aged or above group in China, as one of the most susceptible populations to health misinformation, shares health misinformation on social media. Specifically, informed by the stimulus-organism-response (S-O-R) model (Mehrabian & Russell, 1974) and the cognitive load theory (Sweller, 2011), we bridged the pathway from social media health information seeking to health misinformation sharing on social media. A serial mediation model was proposed to investigate how two psychological factors (i.e., health information overload and health misperceptions) channel the effect of information seeking on misinformation sharing. Additionally, we examined how the need for cognition (NFC), as a critical information processing predisposition, may condition the serial mediation process. The moderated serial mediation model not only unravels the motivators of health misinformation sharing against the backdrop of turning to social media for health information but also sheds light on potential remedies to curb health misinformation sharing.

This study adds to the existing scholarship in three main aspects. Firstly, unlike previous efforts that merely explore the spread of certain types of health misinformation (e.g., COVID-19-related misinformation, see Ahmed & Rasul, 2022), our research focuses on the broad health misinformation without specifying the topic or period, rendering a strengthened external validity. Secondly, since limited knowledge is known about what propels health misinformation proliferation and relevant studies were predominantly conducted in Western societies (Apuke & Omar, 2021), our mechanism based on Chinese survey data broadens the current research scope and enriches the existing explanatory framework. Thirdly, although growing evidence shows that the middle-aged or above group is more vulnerable to health misinformation than other age groups, scholarly attention has been disproportionately allocated to the younger generation (Guan, 2019). Thus, our attempts help to illustrate the psychological process behind sharing health misinformation among this underexamined age group, which is particularly crucial for countries with expanding aging populations like China.

## 2. Theoretical Background

As an updated framework of the S-R (stimulus-response) model, the S-O-R model highlights the role of the organism, which refers to the internal processing of external stimuli (Mehrabian & Russell, 1974). Specifically, the “S” represents the external or environmental cues, such as news exposure (Xiao & Su, 2023). The “O” stands for cognitive and affective processes activated by the stimuli, such as information overload (Zheng et al., 2022). The “R” indicates the final response, mainly comprising behaviors like misinformation sharing (Wu, 2022). Essentially, the S-O-R model emphasizes psychological processing in mediating the relationship between environmental stimuli and behavioral responses.

Recent years have witnessed a growth in adopting the S-O-R model to understand health issues. For instance, Zheng and colleagues (2023) found that seeking health information through different online sources (S) influenced online information overload and trust (O), which further led to cyberchondria (R). Similarly, in a study on how seeking online vaccine information (S) is associated with vaccination intention (R), perceived vaccine information overload, vaccine risk perception, and negative affective response were incorporated as mediators (O), reflecting information seekers’ internal states after information seeking (Zheng et al., 2022). More specifically, in the context of health misinformation sharing, Wu (2022) disclosed that social media dependencies (S) influenced users’ cognitive and affective states (O), ultimately leading to the sharing of COVID-19 misinformation (R). Another work linking the pathway between social media health information seeking (S) and health misinformation sharing (R) confirmed the mediating role of misperception (O) (Tang et al., 2023). These studies illustrate the power of the S-O-R model in explaining health-related behaviors.

Drawing upon the S-O-R model, this study zooms into the organism part by examining the roles of health information overload and misperceptions. The cognitive load theory suggests a ceiling exists on people’s ability to process information, and information overload happens when the amount of available information transcends the ceiling (Sweller, 2011). Although information overload has received scholarly attention in misinformation research (Apuke et al., 2022; Laato et al., 2020; Wu & Pei, 2022), only a few studies have thoroughly disentangled the relationships between online information seeking, information overload, and misinformation sharing. Nevertheless, social media affords an overabundance of information, making it easy to trigger information overload (Laato et al., 2020; Zheng et al., 2022), which poses significant challenges to information seekers. Therefore, it is pivotal to empirically test how seeking health information on social media affects health information overload and misinformation dissemination.

Furthermore, the consequences of health information overload have not been fully recognized and deserve further inquiry in the misinformation environment. Theoretically, information overload represents a suboptimal mental state induced by a heavy load of information (Jiang & Beaudoin, 2016). This mental state, in turn, slows down people’s ability to identify valid information (Muhammed T & Mathew, 2022), makes them recoil from seeking or digesting information (Heiss et al., 2023; Jiang & Beaudoin, 2016; Zheng et al., 2022), and results in inaccurate beliefs and misinformed decisions (Laato et al., 2020). Conceptualized as “beliefs about factual matters are not supported by clear evidence and expert opinion” (Nyhan & Reifler, 2010, p. 305), misperceptions have drawn particular attention in misinformation studies because “people who generate or spread misinformation must hold misperceptions of an issue beforehand” (Su et al., 2022b, p. 2). However, to the best of our knowledge, studies have yet to examine how information overload may impact misperceptions, even though misperceptions are among the possible outcomes of information overload. Our study hence extends the thread by delving into how health information overload relates to health misperceptions, which benefits a nuanced understanding of information overload’s roles in the face of misinformation.

Besides unpacking the organism component, this study further explores how an individual’s information processing predisposition works in the above psychological process. Information processing predispositions are inherent human traits that may shape the relationship between social media information seeking and misinformation-related behaviors (Wu et al., 2023). Thus far, their roles in influencing individuals’ susceptibility to health misinformation have not been fully understood (Tang et al., 2023; Wu et al., 2023). Hence, we focus on NFC, an important information processing style representing how much an individual engages in and enjoys critical thinking (Cacioppo & Petty, 1982), to unravel its function in health misinformation sharing, especially how it may restrain the proliferation of health falsehoods.

Taken together, the overarching theoretical framework integrates the S-O-R model, cognitive load theory, and information processing predispositions. The detailed rationales underlying the relationships are elucidated hereunder.

## 3. Research Model and Hypotheses Development

### 3.1 The direct relationship between social media health information seeking (“S”) and health misinformation sharing (“R”)

Social media have always been criticized as Petri dishes of misinformation and it is inevitable to encounter health misinformation when searching for health information on social media. Several reasons account for this blame. Firstly, the incomplete gatekeeping and fact-checking systems on social media leave chances for the generation and propagation of misinformation (Hameleers et al., 2020; Melki et al., 2023). Secondly, the extensively adopted social curation and algorithmic curation techniques on social media submerge users in an environment rife with attitude-consistent information, heightening the likelihood of misinformation exposure and false belief reinforcement (Lee et al., 2023; Wang & Jacobson, 2023). Thirdly, misinformation has been wrapped as more eye-catching and interesting than authentic information on social media, which easily attracts users’ attention and goes viral (Chen et al., 2015; Kim et al., 2023; Su et al., 2022a). Those factors, coupled with social media’s rich interactivity affordances (e.g., sharing, liking, and hashtags), enable individuals to disseminate misinformation effortlessly on various social media platforms.

Regarding the specific association between social media information seeking and misinformation sharing intention, studies following the uses and gratifications approach revealed that the two components are positively related. For instance, in the COVID-19 context, Apuke and Omar (2020; 2021) found that information seeking, as one of the most important gratifications of using social media, was positively associated with COVID-19 misinformation sharing. Similarly, Chen and Sin (2013) pointed out that information seeking was the primary driving force behind sharing misinformation on social media. Considering the prevalence of misinformation on social media and the tight connection between social media information seeking and misinformation sharing, we believe those phenomena also apply to health (mis)information and are especially prominent for the middle-aged or above population without sufficient ability to discern misinformation. Thus, we posit the following:

H1: Social media health information seeking would be positively associated with health misinformation sharing intention.

### 3.2 Health information overload and misperceptions as mediators (“O”)

Information overload is a critical concept in information processing and is particularly prevalent in the social media era when message recipients cannot fully assimilate or digest the overwhelming online information torrents (Jiang, 2022; Jiang & Beaudoin, 2016). Social media health information seeking leads to health information overload in two ways, namely, quantitatively and qualitatively. Regarding the quantitative side, when searching for health information on social media, it is inevitable to encounter a considerable amount of irrelevant information because search results are often ranked by popularity or algorithms rather than mere relevance (Jiang & Beaudoin, 2016). The superfluous information can be cognitively demanding and burdensome for information seekers. Regarding the qualitative side, health information on social media is a mixture of medical jargon and personal anecdotes, as well as credible information and unverified claims, which makes it difficult for information seekers to navigate the ambiguous, contradictory, and complex information environment (Jiang & Beaudoin, 2016; Zheng et al., 2022).

Additionally, health information overload may lead to health misinformation sharing. Apuke and associates (2022) argued that once individuals feel stressed by processing an influx of information, their motivation to verify the information decreases, which increases the probability of sharing misinformation. The sharing without sufficient deliberation hypothesis got supported by a survey conducted among Nigerian social media users, which revealed that social media information overload was positively associated with misinformation sharing (Apuke et al., 2022). Similarly, in the COVID-19 context, Wu and Pei (2022) discovered that social media information overload was positively associated with individuals’ health anxiety and exhaustion, which were further related to health misinformation sharing. In light of the above rationales, the following hypothesis is proposed.

H2: Health information overload would mediate the relationship between social media health information seeking and health misinformation sharing intention, such that social media information seeking would be positively associated with health information overload (H2a), which would be further positively associated with health misinformation sharing intention (H2b). In terms of the second component of the organism, misperceptions have always been adopted as a mediator in health misinformation studies (Borah et al., 2022; Tang et al., 2023; Xiao, 2022). The existing literature demonstrates that due to the abundance of health misinformation on social media, it is easy for social media users to develop incorrect beliefs by either actively seeking information (Allington et al., 2021; Tang et al., 2023) or coming across information (Borah et al., 2022; Xiao & Su, 2023) on diverse platforms. The essential reason is that social media users are likely to internalize health misinformation in an environment inundated with misinformation, facilitating the crystallization of inaccurate perceptions (Xiao & Su, 2023). Additionally, scholars indicated that since social media intensify selective exposure, it is more frequent for social media information seekers to embrace information that aligns with their preexisting beliefs and disfavor belief-inconsistent information (Tang et al., 2023; Wu et al., 2022). This confirmation bias aggravates misperceptions, given that the original belief may be inaccurate and lacks scientific basis. Moreover, misperceptions are especially prominent among the middle-aged or above group because they underperform in critical thinking and analytical reasoning, making them more receptive to online misinformation (Greene & Murphy, 2020; Xiao, 2022).

Regarding the association between misperceptions and misinformation sharing, ample evidence shows that individuals are likely to share content that they perceive as credible (Valenzuela et al., 2019; Yang et al., 2022). In other words, if social media information seekers deem certain health information reliable, they would hold a positive attitude toward it and are inclined to disseminate it to others. Besides, information sharing has become a common strategy to cope with potential threats and uncertainties (Lu et al., 2022; Tang & Zou, 2021). Therefore, due to the urgency and uncertainty surrounding health issues, social media users may proactively propagate health information they perceive as trustworthy without thoroughly verifying it, leaving chances for the spread of health misinformation (Tang et al., 2023). The above arguments motivate the following hypothesis.

H3: Health misperceptions would mediate the relationship between social media health information seeking and health misinformation sharing intention, such that social media information seeking would be positively associated with health misperceptions (H3a), which would be further positively associated with health misinformation sharing intention (H3b).

Besides the separate mediating roles of health information overload and health misperceptions, we further postulate that information overload serves as an antecedent of misperceptions. According to Tandoc and Kim (2023), perceived information overload hampers individuals from evaluating social media information in depth, leading to analysis paralysis. The paralysis, in turn, promotes news avoidance and results in beliefs in COVID-19 misinformation (Tandoc & Kim, 2023). Similarly, Jiang (2022) found that social media users experience fatigue and exhaustion when suffering from information overload, which hinders them from performing fact-checking. The deficient fact-checking motivation provides an opportunity for developing misperceptions, as people may be unable to differentiate between authentic information and misinformation on their own. In addition, the cognitive load theory posits that excessive information induces decreased cognitive performance and increased judgment errors (Sweller, 2011), which means it could be challenging for recipients to process information accurately with finite mental resources in the face of the information deluge. Therefore, information overload may contribute to misinterpretation, heightening the likelihood of misperception formation.

H4: Health information overload would be positively associated with health misperceptions.

Taking the above hypotheses together, we further put forward the following serial mediation process.

H5: Health information overload and misperceptions would mediate the relationship between social media health information seeking and health misinformation sharing intention serially.

### 3.3 The moderating role of NFC

NFC reflects the rational system when processing information, featuring intentional, conscious, analytical, and affect-free thinking after information exposure (Epstein et al., 1996). According to Austin and colleagues (2016), NFC-oriented individuals tend to process information thoughtfully and systematically rather than relying on heuristic cues. This tendency, in turn, enables critical thinking and results in a skeptical view of persuasive messages (i.e., persuasion resistance) (Austin et al., 2016). Borah (2022) further suggested that high-NFC individuals would carefully monitor information, making them less susceptible to misinformation.

Although NFC has been claimed to have an edge in shielding the adverse effects of misinformation and buffering the effectiveness of persuasive messages, competing empirical findings emerge regarding the moderating role of NFC in the health misinformation setting. For instance, Su and associates (2021) uncovered that the negative indirect effect of international social media use on COVID-19 conspiracy theory endorsement was stronger among respondents with a high level of NFC. In other words, the high-NFC respondents were more likely to discern COVID-19 misinformation in the highly diversified international social media environment and less likely to endorse conspiracy theories. Conversely, Wu and colleagues (2023) disclosed that the positive association between social media health information reliance and health misinformation belief was stronger among the high-NFC group than the low-NFC group. Another study by Tang and colleagues (2023) failed to observe a significant moderating effect of NFC on the relationship between social media health information seeking and health misinformation sharing. Therefore, whether NFC plays a perceptual filter role in the face of health (mis)information remains debatable, leading to the following research question.

RQ1: Would NFC moderate the effects of social media health information seeking on health information overload (RQ1a), health misperceptions (RQ1b), and health misinformation sharing intention (RQ1c)?

Based on the above, the conceptual model of our study is exhibited in Figure 1 with hypotheses and questions indicated.

**Figure 1.**
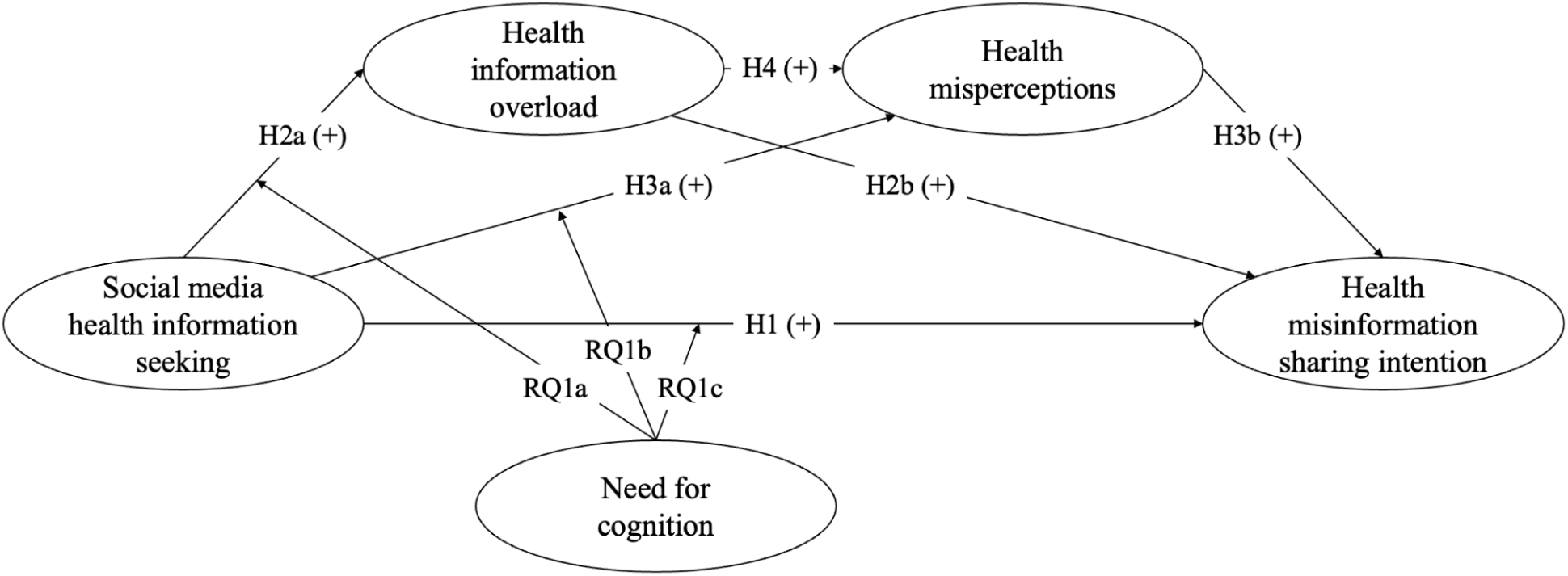
The conceptual model of the current study.

## 4. Methods

### 4.1 Sample

Data were collected through an online survey conducted in mainland China between 1 and 14 February 2023. We followed a volunteer sampling approach due to the deficiency of sampling frames for China’s middle-aged or above group. The questionnaire was administered via *SoJump*, a widely employed survey platform in China. Respondents were first invited to read an informed consent and indicate their willingness to take the survey. Those who agreed to proceed were directed to the formal questionnaire. A total of 500 respondents completed the questionnaire. After excluding those who did not meet the age requirement (i.e., under 45 years old^1^, *N* = 107) and failed to pass the attention check question (*N* = 5), we obtained a final sample of 388 respondents. Among them, the average age was 54.01 with a range from 45 to 74. Slightly more female (53.61%) than male (46.39%) respondents took the survey. The median monthly household income per person ranged from 3K to 6K CNY.

### 4.2 Measures

Prior to the formal survey, we pretested the questionnaire with four Ph.D. candidates majoring in social sciences to ensure the comprehensibility of the wording, and some adjustments were made as per their feedback. Unless indicated otherwise, a 5-point Likert scale ranging from 1 (strongly disagree) to 5 (strongly agree) was employed to measure each variable.

#### Social media health information seeking

In line with previous studies (Ho et al., 2020; Tang et al., 2023; Valenzuela et al., 2019), a single item (i.e., “How often do you seek health information from social media (Weibo, WeChat, Douyin, etc.)?”) was adopted to measure respondents’ health information seeking frequency on social media platforms. A higher score indicates a higher frequency of social media health information seeking (1 = “never”, 5 = “always”; *M* = 3.85, SD = 1.04).

#### Health information overload

Four items adapted from previous studies (Jiang, 2022; Karr-Wisniewski & Lu, 2010; Zheng & Jiang, 2022) were adopted to assess respondents’ health information overload. A sample item was “I receive too much health information every day, making it hard to digest.” A higher score represents greater health information overload (*M* = 3.86, *SD* = 0.77, *α* = 0.82).

#### Health misperceptions

Similar to extant misinformation research (Tang et al., 2023; Valenzuela et al., 2019; Xiao & Su, 2023), we selected seven pieces of widely spread health misinformation based on annual reports released by authoritative institutions in China (e.g., the China Association for Science and Technology) and asked the respondents to indicate how much they agreed with those claims. A sample item was “Fire treatment can cure diseases.” A higher value represents a deeper belief in health misinformation (*M* = 3.58, *SD* = 0.77, *α* = 0.85).

#### Health misinformation sharing intention

Consistent with former studies (e.g., Xiao & Su, 2023), respondents were asked to indicate how likely they were to share the aforementioned seven health misinformation pieces on social media platforms. The higher the score, the more likely to share (*M* = 3.62, *SD* = 0.84, *α* = 0.88).

#### NFC

The six-item version of the NFC scale developed by de Holanda Coelho et al. (2020) was adopted to measure respondents’ NFC levels. Consistent with previous practices (Tang et al., 2023; Wu et al., 2023), two reversed items in the scale were removed to elevate reliability, leaving four items. A sample item was “I would prefer complex to simple problems.” A higher value demonstrated a stronger tendency to engage in and enjoy thinking (*M* = 3.59, *SD* = 0.83, *α* = 0.81).

#### Covariates

Demographic predictors of susceptibility to health misinformation summarized in Nan and colleagues’ (2022) systematic review were incorporated as controls, including age (*M* = 54.01, *SD* = 5.88), gender (53.61% female respondents), educational level (1 = primary school or below, 4 = college or above, *M* = 3.01, *SD* = 0.92), income (1 = below 3K CNY, 6 = above 15K CNY, *M* = 3.10, *SD* = 1.72), ethnicity (25.00% from ethnic minorities), and geographic region (30.15% from rural areas). Besides, religious belief (18.30% had religious beliefs) and personal health status (1 = poor, 4 = excellent, *M* = 2.71, *SD* = 0.87) were also considered since previous studies have suggested their influences on health misinformation-related behaviors (Lee Rogers & Powe, 2022; Sun et al., 2022).

Pearson’s zero-order correlations among all variables are summarized in Table 1.

**Table 1.** Pearson’s zero-order correlations across all variables.

|  | 1 | 2 | 3 | 4 | 5 | 6 | 7 | 8 | 9 | 10 | 11 | 12 |
| --- | --- | --- | --- | --- | --- | --- | --- | --- | --- | --- | --- | --- |
| 1. Age | - |  |  |  |  |  |  |  |  |  |  |  |
| 2. Gender | 0.123* | - |  |  |  |  |  |  |  |  |  |  |
| 3. Education | -0.063 | -0.028 | - |  |  |  |  |  |  |  |  |  |
| 4. Income | 0.168** | 0.162** | 0.021 | - |  |  |  |  |  |  |  |  |
| 5. Ethnicity | 0.200*** | 0.095 | -0.133** | 0.079 | - |  |  |  |  |  |  |  |
| 6. Geographic region | -0.129* | -0.053 | 0.077 | -0.045 | -0.113* | - |  |  |  |  |  |  |
| 7. Religious belief | 0.021 | -0.013 | -0.097 | 0.071 | 0.050 | 0.020 | - |  |  |  |  |  |
| 8. Health status | -0.007 | 0.023 | -0.040 | 0.069 | -0.031 | -0.015 | 0.144** | - |  |  |  |  |
| 9. Social media health information seeking | 0.015 | 0.057 | 0.001 | 0.093 | 0.007 | -0.098 | 0.022 | 0.063 | - |  |  |  |
| 10. Health information overload | 0.058 | 0.066 | -0.096 | 0.100* | 0.087 | -0.080 | 0.017 | -0.028 | 0.226*** | - |  |  |
| 11. Health misperceptions | 0.181*** | 0.119* | -0.157** | 0.186*** | 0.303*** | -0.211*** | 0.160** | 0.070 | 0.298*** | 0.492*** | - |  |
| 12. Need for cognition | 0.193*** | 0.180*** | -0.121* | 0.161** | 0.278*** | -0.124* | 0.091 | 0.041 | 0.240*** | 0.394*** | 0.588*** | - |
| 13. Health misinformation sharing intention | 0.162** | 0.097 | -0.113* | 0.129* | 0.309*** | -0.127* | 0.146** | 0.061 | 0.294*** | 0.477*** | 0.781*** | 0.513*** |

Note. \* $p < 0.05$ ; \*\* $p < 0.01$ ; \*\*\* $p < 0.001$ .

### 4.3 Analytical strategy

Structural equation modeling (SEM) was adopted to analyze the data due to the mixture of latent (e.g., health information overload) and observed (e.g., social media health information seeking frequency) variables in our study. In line with extant studies (Apuke & Omar, 2021; Ho et al., 2022), we first checked the common method bias and collinearity to minimize estimation errors. Secondly, the measurement model was specified using a confirmatory factor analysis (CFA) to ensure the robustness of the measurement component. Thirdly, the structural model was tested to examine the proposed relationships among constructs of interest. All analyses were performed using Stata 17.0. One thing to be noted is that our model is a moderated serial mediation model; therefore, the serial mediation model was examined before the complete model.

## 5. Results

### 5.1 Testing common method bias and collinearity

Common method bias was examined first since all data were gleaned from the same round. Firstly, Harman’s single-factor analysis demonstrated that a single factor explained 37.71% of the total variance, which fell below the threshold of 50% (Dupuis et al., 2017). Secondly, the maximum value in the correlation matrix among the main constructs was 0.78, less than the threshold of 0.90. These results revealed that common method bias was not likely to be a threat (Apuke & Omar, 2021). Regarding collinearity, the maximum value of the variance inflation factor was 2.16, suggesting that collinearity was not a concern in the current study.

### 5.2 The measurement model

Table 2 exhibits the measurement model’s internal reliability, composite reliability, and convergent validity. According to Xiao and Su (2023), most factor loadings were above 0.70, indicating sufficient internal reliability. Additionally, all composite reliability coefficients were greater than 0.80, suggesting satisfactory composite reliability. Regarding convergent validity, all average variance extracted values were above 0.50, implying good convergent validity.

**Table 2.** Construct reliability and validity.

| Construct | Indicator | Factor loading | Cronbach's alpha | CR | AVE |
| --- | --- | --- | --- | --- | --- |
| Health information overload (HIO) | HIO1 | 0.78 | 0.82 | 0.88 | 0.64 |
|  | HIO2 | 0.79 |  |  |  |
|  | HIO3 | 0.82 |  |  |  |
|  | HIO4 | 0.83 |  |  |  |
| Health misperceptions (HM) | HM1 | 0.67 | 0.85 | 0.88 | 0.52 |
|  | HM2 | 0.77 |  |  |  |
|  | HM3 | 0.74 |  |  |  |
|  | HM4 | 0.72 |  |  |  |
|  | HM5 | 0.74 |  |  |  |
|  | HM6 | 0.75 |  |  |  |
|  | HM7 | 0.66 |  |  |  |
| Health misinformation sharing intention (HMS) | HMS1 | 0.71 | 0.88 | 0.91 | 0.58 |
|  | HMS2 | 0.79 |  |  |  |
|  | HMS3 | 0.82 |  |  |  |
|  | HMS4 | 0.78 |  |  |  |
|  | HMS5 | 0.77 |  |  |  |
|  | HMS6 | 0.75 |  |  |  |
|  | HMS7 | 0.74 |  |  |  |
| Need for cognition (NFC) | NFC1 | 0.84 | 0.81 | 0.88 | 0.64 |
|  | NFC2 | 0.84 |  |  |  |
|  | NFC3 | 0.73 |  |  |  |
|  | NFC4 | 0.80 |  |  |  |

*Note.* CR = Composite Reliability. AVE = Average Variance Extracted. The prefix of each indicator is the abbreviation of the corresponding construct.

Furthermore, the Heterotrait-Monotrait (HTMT) ratio was adopted to test the discriminant validity (Xiao & Su, 2023). The maximum value was 0.70, lower than the cutoff point of 0.85, demonstrating a robust discriminant validity of all measures. The overall measurement model also attained a satisfactory goodness-of-fit (*χ*^2^ = 415.04, *df* = 203, *p* < 0.001; *χ*^2^/*df* = 2.04; CFI = 0.95, TLI = 0.94, RMSEA = 0.05, SRMR = 0.04) (Gil de Zúñiga et al., 2022).

### 5.3 The structural model

The serial mediation model was examined first. Consistent with the goodness-of-fit criteria adopted by Gil de Zúñiga and associates (2022), the mediation model fitted the data well (*χ*^2^ = 578.14, *df* = 283, *p* < 0.001; *χ*^2^/*df* = 2.04; CFI = 0.91, TLI = 0.90, RMSEA = 0.05, SRMR = 0.07). All paths were significant (from social media health information seeking to health information overload: β = 0.25, *SE* = 0.05, *p* < 0.001; from social media health information seeking to misperceptions: *β* = 0.19, *SE* = 0.05, *p* < 0.001; from health information overload to health misperceptions: *β* = 0.53, *SE* = 0.05, *p* < 0.001; from misperceptions to misinformation sharing intention: *β* = 0.85, *SE* = 0.04, *p* < 0.001) except the effect of social media health information seeking on health misinformation sharing intention (*β* = 0.03, *SE* = 0.04, *p* = 0.35) and the effect of information overload on misinformation sharing intention (*β* = 0.06, *SE* = 0.07, *p* = 0.35). Accordingly, the mediation process through misperceptions was significant (*effect* = 0.16, *SE* = 0.04, *p* < 0.001, 95% *CI* = [0.08, 0.24]) but the one through information overload was not (*effect* = 0.02, *SE* = 0.02, *p* = 0.36, 95% *CI* = [-0.02, 0.05]). Finally, the serial mediation process was significant (*effect* = 0.11, *SE* = 0.03, *p* < 0.001, 95% *CI* = [0.06, 0.17]).

Figure 2 summarizes the statistical results of the moderated serial mediation model. The model attained a good fit (*χ*^2^ = 572.76, *df* = 313, *p* < 0.001; *χ*^2^/*df* = 1.83; CFI = 0.92, TLI = 0.92, RMSEA = 0.05, SRMR = 0.05) and all factor loadings were significant. The model explained 17.48% of the variance in health information overload, 51.24% of the variance in health misperceptions, and 83.06% of the variance in health misinformation sharing intention.

**Figure 2.**
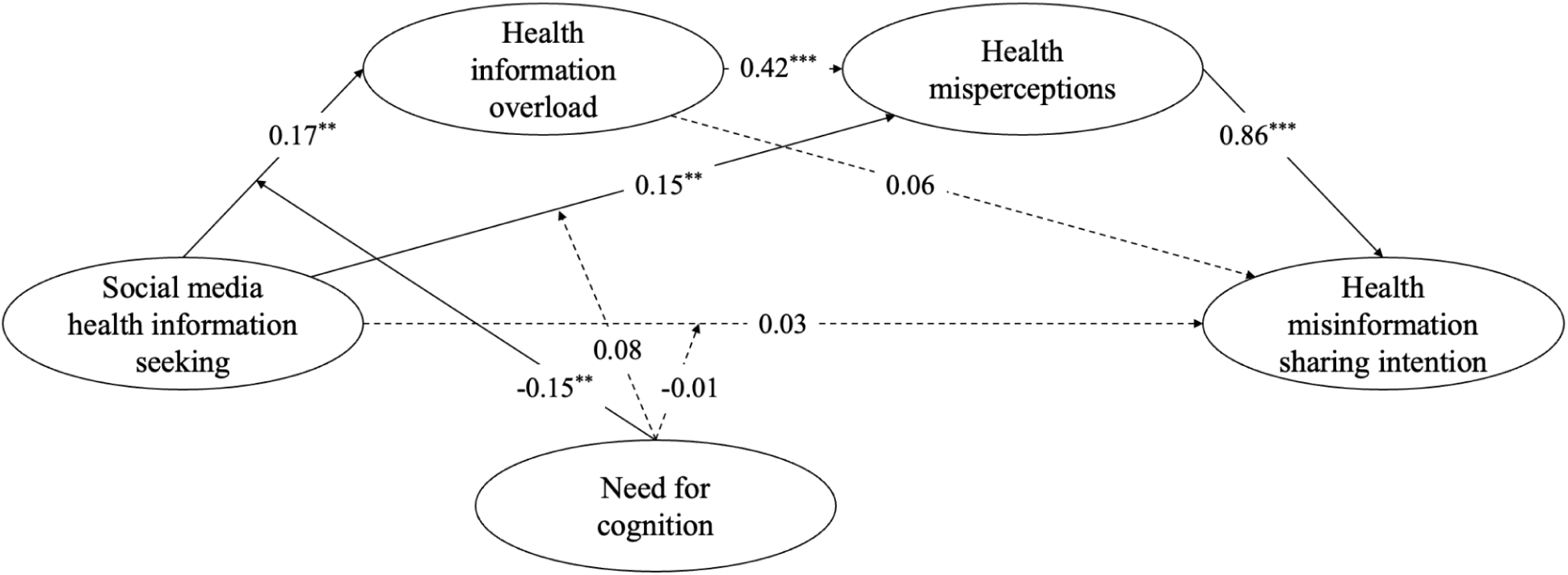
Structural equation model with path coefficients. *Note*. *N* = 388. Standardized coefficients are reported. Control variables are omitted for brevity. Solid lines denote statistically significant paths, whereas dotted lines denote statistically nonsignificant paths. * *p* < 0.05; ** *p* < 0.01; *** *p* < 0.001.

As shown in Figure 2, the association between social media health information seeking and health misinformation sharing intention was not significant (*β* = 0.03, *SE* = 0.04, *p* = 0.39). Thus, H1 failed to receive support.

H2 hypothesizes a mediation process through health information overload. Social media health information seeking was positively associated with health information overload (*β* = 0.17, *SE* = 0.05, *p* < 0.01), supporting H2a. However, the relationship between information overload and health misinformation sharing intention was not significant (*β* = 0.06, *SE* = 0.06, *p* = 0.34), rejecting H2b. The mediation effect was also not significant (*effect* = 0.01, *SE* = 0.01, *p* = 0.36, 95% *CI* = [-0.01, 0.03]). Therefore, H2 was partially supported.

H3 posits a mediation process channeled by health misperceptions. As displayed, social media health information seeking was positively associated with misperceptions (*β* = 0.15, *SE* = 0.04, *p* < 0.01), which in turn was positively associated with health misinformation sharing (*β* = 0.86, *SE* = 0.05, *p* < 0.001). The mediation process was significant (*effect* = 0.13, *SE* = 0.04, *p* < 0.01, 95% *CI* = [0.05, 0.20]). Hence, H3 was fully supported.

H4 postulates a positive association between health information overload and health misperceptions, which is consistent with the statistical results (*β* = 0.42, *SE* = 0.05, *p* < 0.001). Besides, the results lent support to the serial mediation process through health information overload and misperceptions (i.e., H5) (*effect* = 0.06, *SE* = 0.02, *p* < 0.01, 95% *CI* = [0.02, 0.10]).

In terms of the moderating effects (i.e., RQ1), NFC failed to moderate the relationships between social media health information seeking and health misperceptions (*β* = 0.08, *SE* = 0.04, *p* = 0.06), as well as health misinformation sharing intention (*β* = -0.01, *SE* = 0.04, *p* = 0.83). However, the interaction effect of NFC and social media health information seeking on health information overload was significant (*β* = -0.15, *SE* = 0.05, *p* < 0.01). Thus, among respondents with higher NFC, the positive association between health information seeking and information overload was weaker than those with lower NFC.

An additional check was performed to ensure the model’s appropriateness (as displayed in Appendix A^2^). Specifically, we compared the proposed conceptual model to several competing models to determine which fits the data best. The model fit indices revealed that the current model outperformed the alternatives.

## 6. Discussion and Conclusion

Informed by the S-O-R model and the cognitive load theory, this study constructs a conceptual model to illustrate the psychological mechanism behind health misinformation sharing among the middle-aged or above group in China. Facing the onslaught of social media health misinformation, penetrating the underlying mechanism benefits misinformation regulation and targeted interventions. Results demonstrate that seeking health information on social media was indirectly associated with health misinformation sharing through health information overload and misperceptions. Meanwhile, the direct relationship was not significant. Additionally, the correlation between health information overload and misinformation sharing intention was not supported. As a moderator, NFC only buffers the positive association between health information seeking and health information overload. These intriguing findings merit detailed discussion.

Firstly, as indicated in the literature review, our study is among the few to decipher how information overload links to misperceptions in the context of health misinformation. Results bolster the proposed serial mediation process, which parallels prior studies suggesting that health information overload could be induced when social media information seekers need to handle a tremendous amount of health information from multiple sources with uneven qualities (Jiang, 2022; Jiang & Beaudoin, 2016). Additionally, health information overload brings about undesirable consequences, such as mental stress, message fatigue, and inadequate information elaboration (Jia et al., 2023; Jiang, 2022). When information seekers feel exhausted in processing health information, their judgment accuracy is likely to be impaired, leading to misinterpretation and misperception (Khaleel et al., 2020). As an essential antecedent of misinformation sharing, misperceptions contribute to the final behavioral intention (Su et al., 2022b; Yang et al., 2022). The discovered chain mediation pattern affords a deeper insight into the role of health information overload in health misinformation dissemination, which is particularly concerning for the middle-aged or above group because this group lacks the expertise and necessary digital skills to check the authenticity of online health information (Wu et al., 2022). That is to say, the middle-aged or above population is likely to be stuck in health information overload after seeking health information on social media due to their inability to sort through the voluminous information. The overload, in turn, would likely develop into misperceptions and health falsehood sharing intention because of the insufficient digital literacy of this group.

Secondly, the mediation path through health misperceptions differs from the one through health information overload. Regarding the first halves of the two paths, the regression coefficients of social media health information seeking were nearly identical (i.e., *β* = 0.15 vs. *β* = 0.17). The findings align well with previous studies illustrating that overwhelming online health information could burden an information seeker’s cognitive system (e.g., Zheng et al., 2022; Zheng & Jiang, 2022). Meanwhile, the rampant health misinformation on social media can also facilitate the internalization of misinformation since it is difficult for information seekers to circumvent misinformation when seeking health information online (Tang et al., 2023). Therefore, the positive relationship between social media health information seeking and health misperceptions is unsurprising.

Whereas the first halves of the two paths were similar, the second halves differed in terms of the nonsignificant association between health information overload and health misinformation sharing intention. Combined with the serial mediation process, a possible reason is that as a mental state engendered by the imbalance between excessive information and limited processing capacity (Eppler & Mengis, 2004), information overload needs to influence the belief session before translating into behavioral intentions. This finding is not without scholarly support. For instance, Wu and Pei (2022) suggested that health anxiety and exhaustion serve as mediators between information overload and health misinformation sharing. Moreover, this finding somewhat challenges one prior research that found a significant relationship between information overload and misinformation sharing behavior (Apuke et al., 2022). A plausible explanation is that Apuke and associates (2022) targeted general social media information, while our study focused solely on health information on social media. One notable difference between the two types of information is that health information is more personally relevant, which means adopting the information or not may have immediate and direct impacts on the recipients (Ma et al., 2023). As a result, even when experiencing health information overload, the middle-aged or above population may be cautious and prudent when making the sharing decision, as sharing inaccurate or inappropriate health information could harm others’ well-being and interpersonal relationships. Only when unverified health information derived from information overload has been integrated into the belief system would information overload affects health misinformation sharing intention. The nonsignificant mediation process through health information overload lends further credence to the necessity to delve deeper into the organism part for a more nuanced understanding of health misinformation sharing.

Thirdly, despite NFC’s facilitating effect on critical thinking and elaborative information processing (Austin et al., 2016; Borah, 2022), it only moderated the relationship between social media health information seeking and health information overload. This suggests that NFC acts as a mental filter for screening the sheer amount of health information available on social media. However, when it goes beyond the mental state part (i.e., health information overload) and comes to the perceptual or behavioral part (i.e., health misperceptions and health misinformation sharing), NFC’s role is rather limited. For one thing, health misperceptions are formed over a long period and are resistant to change, which is especially true for the middle-aged or above group with relatively richer life experiences. Previous studies have also shown that health misperceptions were positively associated with long-lasting general misperceptions (e.g., Borah et al., 2022). Hence, health misperceptions among the middle-aged or above population may be more stable, deep-rooted, and intractable than health information overload, which fall beyond the mitigating effect of NFC. For another, the link between health information seeking and misinformation sharing intention is rather complicated. Studies suggested multiple mediators, such as interpersonal communication and mental reflection, channel the process from information exposure to behaviors (Cho et al., 2009; Shah et al., 2007). Accordingly, the direct effect of social media health information seeking and health misinformation sharing intention is hard to be moderated by a single information processing predisposition. This result reminds researchers to scrutinize the process-oriented mechanism behind information behavior and investigate the specific roles of moderators in each segment.

### 6.1 Theoretical and practical implications

What drives health misinformation sharing is an ongoing issue that requires more fine-grained evidence. This study advances extant misinformation sharing literature by unveiling how health information overload functions in the health misinformation sharing setting. By integrating the S-O-R model and the cognitive load theory, this study bridges health information overload and health misperceptions, which were understudied previously. Notably, the significant serial mediation process and the simple mediation process through health misperceptions reveal that health information overload is not always the direct cause of health misinformation sharing. As a mental state provoked by overwhelming online information, only when it affects the belief system (i.e., health misperceptions) could misinformation sharing intention be triggered. This intricate mechanism affords a nuanced understanding of information overload’s role in the health information field.

Another theoretical contribution pertains to the differential moderating effects of NFC within the proposed mediation process. Although NFC has been touted as an alleviator of misbelief and misinformation-related behaviors, we found that NFC is effective in ameliorating health information overload rather than misperceptions and sharing intentions that are more enduring and tangled. This finding invalidates the simplistic thinking that NFC is a panacea for combating misinformation. Instead, it informs that a subtle perspective is indispensable to better comprehend how NFC works in the middle-aged or above group with a relatively solid belief system.

This study also encapsulates implications for health misinformation intervention. First of all, due to social media health information seeking positively related to both health information overload and health misperceptions, it is urgent for social media platforms to optimize fact-checking and gatekeeping systems to enable timely detection and regulation of health misinformation. For instance, by employing the “related stories” function (i.e., providing matched stories in addition to the original information) to correct potential health falsehoods and reduce misperceptions (Bode & Vraga, 2015). Secondly, since health information overload contributes to health misperception formation, one viable strategy is inviting healthcare professionals and authoritative public health institutions to communicate directly with middle-aged or above health information seekers in cyberspace (Jiang & Beaudoin, 2016; Oktavianus & Bautista, 2023), which would ease their information burdens and help them locate reliable information easily. Thirdly, although NFC only moderates a single path in our mechanism, its shielding role against health misinformation should not be downplayed. Regarding the middle-aged or above group, certain media literacy-enhancing programs may be a silver lining in reinforcing critical thinking skills and the ability to navigate the infosphere. Even though this group may struggle with discerning health misinformation, tailored training may be effective in encouraging them to consume health information from reliable sources, thus reducing the likelihood of information overload.

### 6.2 Limitations and future directions

This study is not free from limitations. Firstly, we only considered health information seeking on social media in general. However, diverse social media platforms offer different affordances that may impact health information overload and misperceptions differently (Oktavianus & Bautista, 2023; Vraga & Bode, 2018). Future studies need to dismantle social media appropriately for a more granular mechanism. Secondly, the organism part is constituted by both cognitive and affective processes (Zheng et al., 2022). The current study only considers the cognitive side but neglects the affective side, necessitating future efforts to supplement the jigsaw puzzle. Thirdly, the cross-sectional nature of this study precludes us from ascertaining causality. Therefore, multi-wave surveys or other longitudinal designs are warranted to pursue a clear causal relationship among the constructs.

## Data Availability

All data produced in the present study are available upon reasonable request to the authors.

## Notes

1. 45 is the starting point of middle age according to the definitions of “middle age” in Merriam-Webster and Oxford English Dictionary.
2. The appendix is available upon reasonable request (by contacting the first author).

